# Stabilizing the return to normal behavior in an epidemic

**DOI:** 10.1101/2023.03.13.23287222

**Authors:** Tyrus Berry, Matthew Ferrari, Timothy Sauer, Steven J. Greybush, Donald Ebeigbe, Andrew J. Whalen, Steven J. Schiff

## Abstract

Predicting the interplay between infectious disease and behavior has been an intractable problem because behavioral response is so varied. We introduce a general framework for feedback between incidence and behavior for an infectious disease. By identifying stable equilibria, we provide policy end-states that are self-managing and self-maintaining. We prove mathematically the existence of two new endemic equilibria depending on the vaccination rate: one in the presence of low vaccination but with reduced societal activity (the “new normal”), and one with return to normal activity but with vaccination rate below that required for disease elimination. This framework allows us to anticipate the long-term consequence of an emerging disease and design a vaccination response that optimizes public health and limits societal consequences.

**Significance Statement:** The experience of the COVID-19 pandemic has revealed that behavior can change dramatically in response to the spread of a disease. This behavioral response impacts disease transmission. Predicting future outcomes requires accounting for the feedback between behavior and transmission. We show that accounting for these feedbacks generates long-term predictions about disease burden and behavior that can guide policy.

---

**T**o understand and control epidemics, models have been developed that reflect the fundamental properties of infectious disease transmission (1). To embody biological understanding and develop effective policy these models rely on abstractions of complicated phenomena: mortality, reinfection, vaccination, loss of immunity and spatial networks (2). Nevertheless, a substantial barrier to progress has been that transmission depends on human behavior, which is impossible to model in detail. To meet this challenge, we must consider all possible responses with minimal assumptions about the behavioral response to disease.

A hallmark of classical models for emergent epidemic dynamics is a large initial outbreak with final size larger than the critical herd immunity threshold (3). The initial emergent epidemic is followed by a period of low prevalence and then outbreaks of much smaller magnitude. This phenomenon raised concerns about the magnitude of the initial waves of infection of Ebola in 2014 (4), SARS-CoV-2 in 2020 (5, 6), and Mpox in 2022 (7, 8), and the potential strain on health systems from such large initial epidemic waves in the absence of behavioral restrictions. However, in all three settings, the initial epidemic wave was curtailed by behavioral change that resulted from a combination of individual behavior to limit risk of exposure and top-down restrictions.

For example, as shown in Fig. 1, in the first year of the SARS-CoV-2 pandemic, before the emergence of the first meaningful immune-escape variant (Alpha in November 2020), many local regions saw a second wave of the original wild-type virus that was equal to, or larger than, the magnitude of the initial emergent epidemic. This implies that behavioral changes, whether individual behavioral or legislated closures, may have limited the size of the first wave, which left a sufficiently large susceptible population that a second wave began when behavior and contact patterns returned towards pre-SARS-CoV-2 levels. The collective experience of these recent global emergence events suggests that disease modeling frameworks that do not account for behavioral change are insufficient to predict the dynamics of the emergence of pathogens exhibiting sufficient morbidity and mortality that will drive behavioral change.

**Fig. 1.**
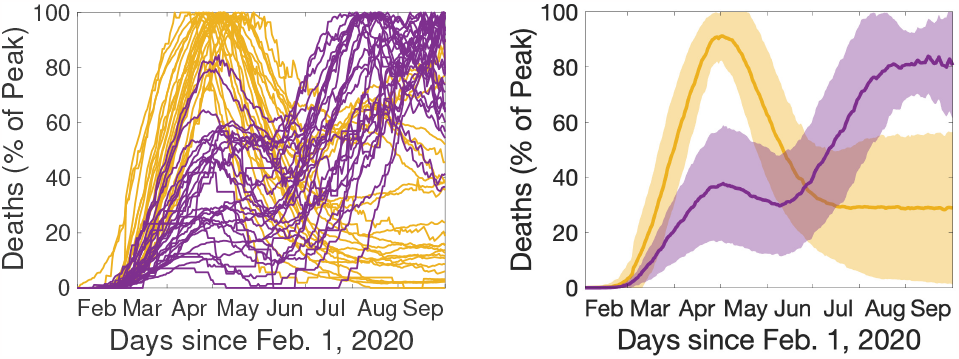
SARS-CoV-2 deaths exhibited multiple increasing waves in some US states prior to documented immune escape. Left: SARS-CoV-2 deaths for 54 US states and territories over the period from Feb. 1, 2020 to Oct. 1, 2020 smoothed with a 56-day moving average and normalized by the peak deaths. Note the large gap in peaks between June and July; the traces cluster into two groups: Peak before June (yellow) and peak after July (purple). Right: Aggregating these two clusters (solid line is the cluster mean and shaded area shows one standard deviation), we see that the yellow cluster exhibits a large initial peak with either no second wave or a smaller second wave (during this time period), whereas the purple cluster exhibits two increasing waves. The dynamic exhibited by the purple cluster cannot be captured with a simple compartmental model and is difficult to explain with spatial dynamics, motivating us to introduce a framework for modeling behavior as a possible explanation. Analysis based on the JHU CSSE COVID-19 Data (15) available at https://github.com/CSSEGISandData/COVID-19.

Behavioral modeling can take many forms depending on whether the behavior patterns of interest are exogenous or endogenous to the disease. Exogenous effects on spread of disease include seasonality or long-established societal patterns of behavior. These are distinguished by a lack of dependence on the state of the disease (number of susceptible, infected, or recovered people) and can be modeled as external covariates; e.g. transmission rate as a function of relative humidity (9) or contact rates as a function of time of year (10). In contrast, endogenous effects (i.e. feedbacks) are dependent on the state of the system (e.g. incidence), including individual choices to modify behavior or policy changes that influence behavior in response to incidence or mortality. Traditional compartmental models omit such feedbacks and are unable to reproduce the breadth of phenomena illustrated in Fig. 1.

Exogenous variables have been used retrospectively to account for observed behavioral phenomena coincident with epidemic dynamics. For example, modern technology such as cell phone based mobility data has enabled exogenous modeling of behavior (11–13). Modeling behavior as a function of exogenous variables permits only retrospective evaluation of the interaction between behavior and transmission. Policy decisions need to anticipate future changes in behavior and thus require a framework that can account for future behavioral change.

In this article we show that the addition of a population level behavioral feedback (between incidence and transmission rate) to the classical SIR model, under a surprisingly weak set of assumptions, implies the existence of three possible equilibrium states: (1) for high vaccination rates, disease eradication, (2) for a medium range of vaccination, an endemic equilibrium with return to normal activity, (3) for low vaccination rates, a “new normal” equilibrium with reduced societal activity. We will also show that the SIR model with activity term can have a wider range of stereotypical behavior, which includes qualitative dynamics during emergence consistent with those shown in Fig. 1.

We show how a wide range of possible endogenous behavioral responses (e.g. distancing, masking, hygiene) can be introduced in a compartmental modeling framework (eg. susceptible, infectious, and recovered, or SIR (14)) in a completely general way. Rather than specify a particular model of behavioral response, we choose reasonable and intuitive properties as assumptions to constrain the form of the behavioral response.

There has been significant work analyzing models with feedback between incidence and vaccination behavior (willingness or hesitancy) (16, 17). Bauch and Earn (16) showed the existence of stable equilibrium vaccination demand that can explain the challenge of attaining universal coverage. There has comparatively little work modeling feedback between incidence and activity (18, 19) as applied to behavioral interventions to limit transmission. Current methods typically rely on choosing a particular model for the feedback (20–27). The key advance here is that we avoid the problematic issue of model specification, so the conclusions we reach are widely applicable, including novel emergence scenarios in unknown behavioral contexts.

Consider a standard disease modeling framework (1, 14) for a single well-mixed population that includes vaccination and loss of immunity. We reflect the endogenous/exogenous dichotomy by decomposing the transmission rate, *β*, into a product of exogenous and endogenous components. The endogenous response is represented with a single variable, *a*, (the instantaneous *activity* of individuals averaged over the population) that quantifies the instantaneous rate of effective behavioral interactions. Rather than specify an exact model for the activity dynamics, we assume that the rate of change of activity is determined by an unspecified *reactivity function*. Without specifying the reactivity function, we base our results on the following three assumptions:

## A1. Reactivity

Change of activity depends on the current level of activity and incidence of infection.

## A2. Resilience

When incidence of infection is zero, activity will return to a baseline level.

## A3. Boundedness

Activity does not exceed the baseline level.

*Reactivity* reflects the assumption that the population chooses its aggregate activity level based on information available; specifically the currently observed activity level and knowledge of disease incidence. This means that the reactivity function, *F*, is a function of activity, *a*, and disease incidence, *c*, or *F* (*a, c*), and does not depend on other variables. Thus, reactivity does not reflect exogenous influences.

We define a *baseline* activity level as the level of activity that the population would go to if the disease were removed and the activity was allowed to stabilize.

*Resilience* is here defined as the ability of the activity to spring back to the previous condition when distorting forces are removed. In this case, new infections are a distorting force, so *resilience* is the assumption that when disease incidence is zero the activity averaged over the population will return towards the baseline level. We also assume that, when there is no incidence, the baseline activity level is stationary.

*Boundedness* asserts that the baseline activity level of the population that exists in the absence of infection is also the maximum activity level. We assign this maximum level to be 1 in arbitrary units, so that the activity level *a* is always between 0 and 1.

Using only these assumptions, we show that the disease equilibria and stability are determined almost entirely by the vaccination rate, *v*, regardless of the behavioral model. We illustrate that accounting for an endogenous behavioral feedback gives rise to a novel equilibrium en route to the classical vaccine-based elimination threshold. The existence of this novel equilibrium can be used as a way-point to guide policy to achieve a return to normal behavior coincident with disease control.

## 1. Results

Starting from the *reactivity* assumption (A1), we first developed a framework for incorporating any reactive behavioral dynamics into the compartmental disease modeling paradigm (see Methods). The state of the modeled disease at any given time can be characterized by three variables (Fig 2): the percentage of the population that is susceptible, *s*, the percentage infected, *i*, and the activity relative to the baseline, *a*. The novelty here is that the reactivity function, *F*, which determines the feedback between activity and infection rate, is left completely unspecified. This means our results will apply very broadly to any behavioral response that satisfies our basic assumptions.

**Fig. 2.**
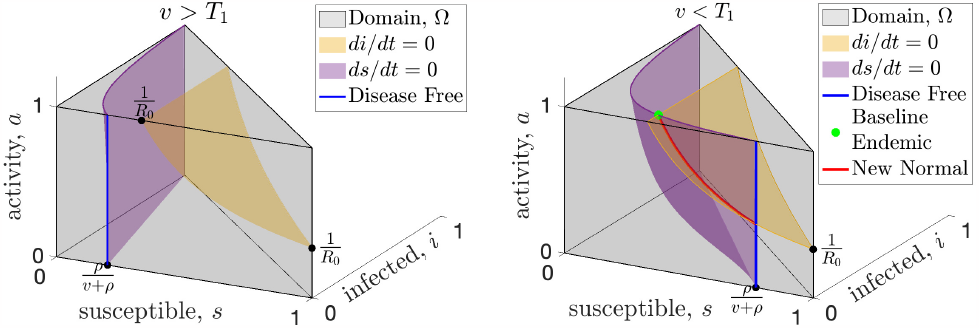
Universal equilibria of resilient behavioral responses with high (left) and low (right) vaccination. The state space (gray shading) of the SIR model with endogenous behavioral feedback plotted on the susceptible (*s*), infected (*i*), and activity (*a*, where *a* = 1 represents the baseline activity level) axes. The susceptible and infected population sizes are instantaneously constant along the purple and yellow surfaces, respectively. An equilibrium must occur along the blue line that shows the intersection of the purple surface with the front of the domain (the *i* = 0 plane) or along the red curve that shows the intersection of these surfaces. When vaccine rates, *v*, are greater than the critical threshold, *T*_1_ (see left panel), the only equilibrium is disease-free (blue) and resilience will drive the activity to baseline which is the top of the blue line. When vaccination rates drop below the *T*_1_ (right panel), the baseline endemic equilibrium (green dot) is created, along with at least one new normal endemic equilibrium which can be anywhere along the red curve.

First, for any model with reactivity (Assumption A1), we find a universal vaccination threshold, *T*_1_, that is independent of the feedback between activity and incidence. When the vaccination rate is above this threshold any equilibrium must be disease-free. Fig. 2 illustrates the surfaces where the infected population (yellow) and the susceptible population (purple) are not changing; an equilibrium can only happen at the intersection of these two surfaces, or where the purple surface intersects the *i* = 0 plane (disease-free). When the vaccination rate is greater than *T*_1_ the only equilibrium is disease-free (Fig. 2a, and Supporting Information figure Fig. S.1a).

Second, by assuming resilience (Assumption A2), we prove that when the vaccination rate is above *T*_1_ the disease-free equilibrium is stable in the face of baseline activity (Fig. S.1a). Resilience assumes that when incidence is zero (disease-free) and activity is below baseline, then activity will increase. While this seems intuitive it does not imply stability by itself. Stability requires that even if we perturb the disease-free equilibrium by introducing a small number of infections, the system must return to the disease-free equilibrium. In Theorem 3 (Supporting Information), we prove that the disease-free equilibrium is in fact stable, as long as the vaccination rate is above *T*_1_.

Assuming both reactivity (*A1*) and resilience (*A2*), when the vaccination rate drops below the universal threshold *T*_1_, the disease-free equilibrium becomes unstable, and endemic equilibria become possible (Fig. 2b). One novel equilibrium, which we call the *baseline endemic equilibrium*, is stable even when activity is at baseline (*a* = 1). For a baseline endemic equilibrium to exist, we only require that normal activity be stationary for this incidence level, meaning that *F* (1, *c*) = 0. Not every reactivity function, *F*, will have a baseline endemic equilibrium, and we give several examples in Section 2 of reactivity functions that show the range of possibilities. If the baseline endemic equilibrium does exist, the infection rate at equilibrium depends on the vaccination rate, but is independent of the behavioral model.

While not as desirable as a disease-free equilibrium, an endemic equilibrium with baseline activity (*a* = 1) may still be preferred to permanently modifying behavior, so it is important to determine its stability. Recall that a bounded behavioral response limits the average activity, *a*, to be at most the baseline level, *a* = 1, by imposing *A1*. In Theorem 4 (Supporting Information), we show that for any bounded behavioral response, there will be a second vaccination threshold, *T*_2_, which determines the stability of the baseline endemic equilibrium. The *T*_2_ threshold is given by (Supporting Information),

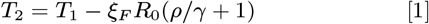

where *ξ*_*F*_ is a constant that depends on the properties of the reactivity function, *F*, near the baseline endemic equilibrium, *R*_0_ is the average number of infections after contact in a fully susceptible population or *basic reproduction number*, and *ρ* and *γ* are rates (see Supporting Information). The *ξ*_*F*_ constant will often be positive, and in these cases the *T*_2_ vaccination threshold will be lower than the *T*_1_ threshold. In these cases, when the vaccination rate is higher than *T*_2_ but less than *T*_1_, the baseline endemic endemic will be stable. For some reactivity functions, the constant *ξ*_*F*_ can be negative or zero, and for these reactivity functions the baseline endemic equilibrium will not be stable for any vaccination rate. Once the reactivity function, *F*, is specified, *T*_2_ can be computed explicitly and we show how to compute *T*_2_ along with several examples in the Supporting Information. This shows that even when the classical threshold for effective vaccination cannot be achieved, there can still be a substantial benefit at a lower vaccination rate. As long as the vaccination rate exceeds the new *T*_2_ threshold, the baseline activity level will be stable (see examples Fig. 3).

**Fig. 3.**
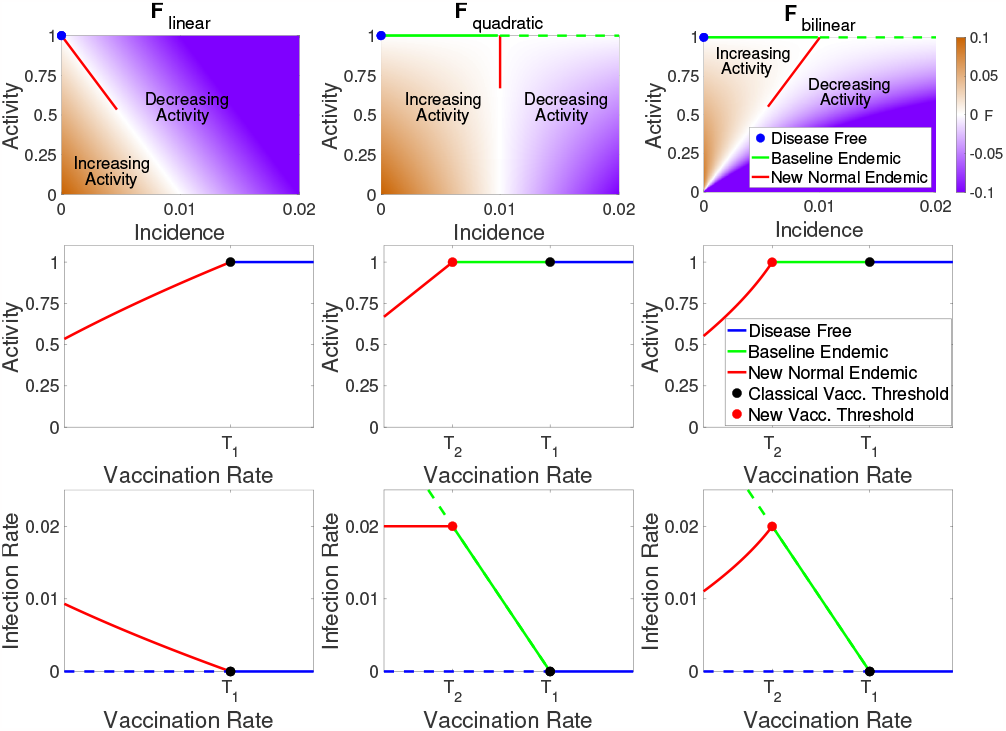
Vaccination may increase or decrease infection rate depending on the form of the behavioral response. Relationship between activity, incidence, and vaccination rate for three example reactivity functions (columns). In the top row we illustrate zones of increasing (brown) and decreasing (purple) activity as a function of incidence; white indicates regions where activity is stationary (at least instantaneously). In the middle row the equilibrium activity is shown as a function of vaccination rate with colors indicating the disease-free, baseline endemic, and new normal regimes. The bottom row indicates the equilibrium incidence as a function of vaccination; stable equilibria are shown as solid lines and unstable equilibria as dashed lines. Note that when vaccination is less than *T*_2_ (the new normal), increased vaccination may lead to either higher (bottom right) or lower (bottom left) infection rates depending on the reactivity function.

When the vaccination rate is below both the *T*_1_ and *T*_2_ thresholds (e.g. early stages of a novel disease before a vaccination, *v* = 0) both the disease-free equilibrium and the baseline endemic are unstable and there is no stable equilibrium with baseline activity. In Theorem 6 (Supporting Information) we prove that there is at least one new equilibrium (Fig. 2b), which we term a “new normal” endemic equilibrium. Unlike the disease-free and baseline endemic equilibrium, the incidence rate at the new normal endemic equilibrium depends on the form of the behavioral feedback and implies long-term behavioral changes with activity level below baseline. When vaccination is below both thresholds, the stability of the new normal endemic cannot be determined universally, and it may have a complicated dependence on the details of the behavioral feedback and exhibit periodic cycles or chaos.

## 2. Examples

We emphasize that our results apply to any reactivity function, *F*, that satisfies A1 - A3. To illustrate our results we introduce three basic examples of reactivity functions.

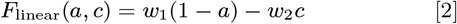

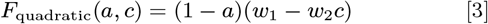

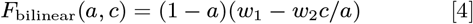

These functions all satisfy resilience and boundedness for any *w*_1_, *w*_2_ > 0 and we illustrate them in the top row of Fig. 3 for *w*_1_ = 0.1 and *w*_2_ = 10. The first two functions (2, 3) are important because they are the leading order approximation of any reactivity function. Note that the function *F*_quadratic_ is quadratic in *a* since *c* = *aBsi* and similarly *F*_bilinear_ is bilinear in *a* and *i*. In the Supporting Information we show that the simplest model, *F*_linear_, does not have a baseline endemic equilibrium because *T*_2_ = *T*_1_. Both *F*_quadratic_ and *F*_bilinear_ have *T*_2_ < *T*_1_, so for vaccination rates between these thresholds the baseline endemic is stable.

The primary difference between the three example reactivity functions is how quickly the equilibrium level of activity falls off as vaccination rate decreases (Fig. 3). For *F*_linear_ the activity versus vaccination curve is concave down, and this moderate response results in a new normal infection rate that increases rapidly as vaccination rate decreases (Fig. 3). For *F*_quadratic_ equilibrium activity increases linearly with vaccination rate. This model has the interesting feature that a decrease in vaccination rate leads to a decrease in activity that maintains a constant level of infection in the new normal endemic. Finally, *F*_bilinear_ has the most robust behavioral response, with a concave up increase in activity as vaccination rate increases. This response results in infection rate increasing as vaccination rates increase. Thus, when initially introducing a vaccination to a population the infection rate may initially increase until the critical vaccination rate threshold *T*_2_ is reached and the baseline endemic is stabilized. This will especially be the case for a new vaccination that is being gradually rolled out, since a slow change in the vaccination rate can help keep the system near equilibrium as the new normal shifts.

Finally, we note a fascinating feature of *F*_bilinear_. If we consider the fraction of the population that remains susceptible to infection as approximately constant and set 1−*a* as ‘distancing’, then we recover a form equivalent to the Lotka-Volterra predator-prey model where infections, *i*, play the role of prey and distancing, 1−*a*, plays the role of the predator (Supporting Information equation Eq. (12)). Oscillations are present at the beginning of the epidemic, when the susceptible population is large and almost constant. The oscillations are not damped, but they have a very slow decay due to the slow decrease in the susceptibles. Finally, the model exhibits a phase transition when the the susceptible population drops below 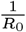, at which point the predatorprey oscillations cease and the system reverts to a more typical epidemic trajectory allowing the system to come to equilibrium. These oscillations depend on the vaccination rate (Fig. 4, Fig. S.3). This illustrates how behavioral feedback can lead to a wide range of epidemic dynamics including oscillations that are independent of any external (e.g. seasonal) forcing.

**Fig. 4.**
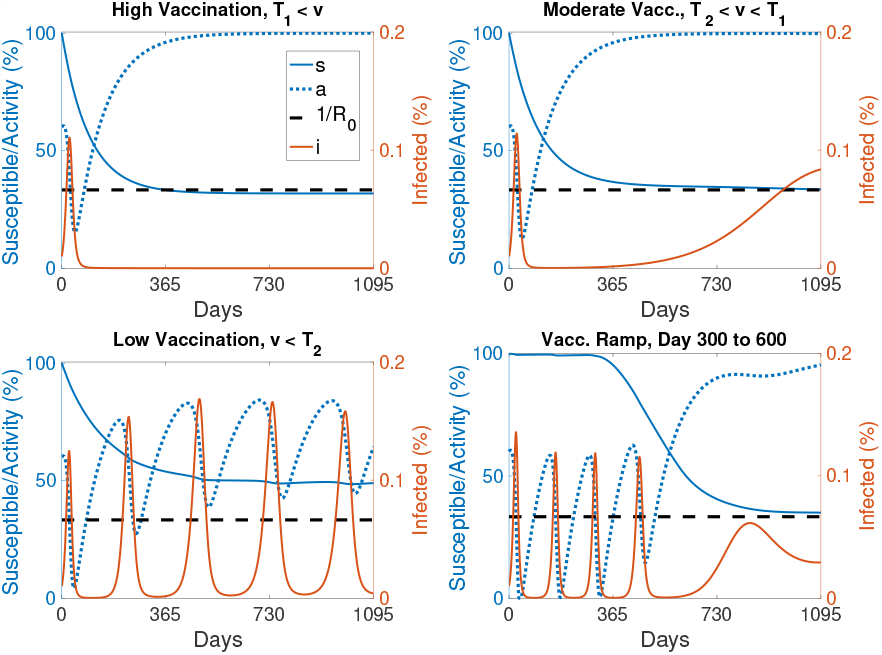
Introducing resilient activity accounts for a wide range of epidemic dynamics. Examples of the dynamics of the reactivity function *F*_bilinear_ with high vaccination (top left, *v* > *T*_1_), moderate vaccination (top right, *v* between *T*_2_ and *T*_1_), and low vaccination (bottom left, *v* < *T*_2_). Finally (bottom right) we simulate 300 days without any vaccination followed by a linear ramp up in vaccination between days 300 and 600 to a fixed moderate vaccination rate after day 600. Susceptible population, *s* (solid blue), activity level, *a* (dotted blue), and the phase transition level 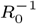 (dashed black) are scaled to the left axis while the infected population, *i* (solid red) is scaled to the right axis. (See equation Eq. (17) in the Supporting Information for details and Fig. S.3 for more examples.)

## 3. Discussion

The collective experience of recent global emergence events suggests that the conventional disease modeling framework is insufficient to predict the dynamics of the emergence of pathogens with severity or mortality that will drive behavioral change. Notably, the standard SIR-model overestimates the expected magnitude of an initial outbreak (in the absence of knowledge about future individual or legislated behavioral change) and consequently underestimates the expected time to and magnitude of subsequent waves. Here we describe the dynamics and equilibria of a novel SIR-type model with a general formulation of behavioral feedbacks.

In the first year of the SARS-CoV-2 pandemic, before the emergence of the first meaningful immune-escape variant (Alpha in November 2020), many places (see Fig. 1) saw a second wave of the original wild-type virus that was equal to, or larger than, the magnitude of the initial emergent epidemic. Behavioral change in response to the initial wave may have left a sufficiently large susceptible population to permit a second, larger wave when behavior and contact patterns returned towards pre-SARS-CoV-2 levels. We find that behavioral feedbacks that reduce contact rates in response to increasing infection incidence can produce these novel dynamics in the transient period of emergence. These behavioral feedbacks also generate novel endemic equilibria characterized by either persistent behavior restriction or a return to pre-emergence behavior levels if vaccination is introduced and sufficiently high.

We have shown that for a broad range of behavioral feedbacks between the incidence of infection and activity that contributes to transmission (e.g. contact rates or hygiene) there exist two novel equilibria in addition to the classic vaccine-based herd immunity threshold. While coordinated behavioral interventions may be sufficient to drive incidence to 0, e.g. as was seen for SARS-CoV-1 in 2004 (28), and Ebola outbreaks (29) prior to the incorporation of vaccination in outbreak response (30), such interventions alone cannot stabilize the disease-free equilibrium if behavior exhibits resilience. In the absence of vaccination there are no stable equilibria that have a return to normal activity. SARS-CoV-1 is the rare example of a pathogen that emerged and was eradicated in the absence of a vaccine; however, reintroduction from an animal reservoir remains possible (31) and the relaxation of the behavioral interventions (28) render the current disease-free state unstable (32).

The newly identified regime with vaccination between *T*_2_ and *T*_1_ has substantial policy implications for emerging infections and eradication. In the absence of vaccines, non-pharmaceutical interventions remain an important part of pandemic response for emerging infections and can be onerous. The SARS-CoV-2 pandemic led to dramatic economic (33) and educational (34, 35) consequences. Planning for a safe return to pre-emergence activity can minimize these off-target effects. While eradication may still be a goal, vaccination at a level *T*_2_ lower than the classic herd immunity threshold *T*_1_ permits a return to pre-emergence activity while maintaining a stable, non-zero incidence. Furthermore, attaining *T*_1_ may be challenging, particularly in the face of vaccine hesitancy, vaccine administration logistics, or uncertainty about the rate of loss of immunity. The existence of *T*_2_ suggests a midpoint goal for vaccination rate that can be used to motivate vaccination efforts.

The existence of the vaccination regime between *T*_2_ and *T*_1_ may further be useful in policies for endemic infections. The only benefits to vaccination in the standard SIR modeling framework without behavioral feedbacks are reductions in morbidity and mortality. This new model implies additional societal change, in the form of the increased activity, that may be stabilized at or above a lower vaccination threshold *T*_2_. Whether this represents a societal benefit or not will be highly epidemic specific. For example, vaccination rates above *T*_2_ may allow for relaxation of pre-screening requirements and the costs inherent in such programs. Alternatively, one could imagine an increase in risky behaviors, e.g. decreased mask usage as vaccination increases. The positive correlation between vaccination rate and equilibrium incidence under the *F*_bilinear_ function could lead to population level assessment of vaccine failure driven by the behavioral response. Any specific predictions of such phenomena is speculative without a mechanistic understanding of the explicit nature of the feedbacks. For example Funk et al. (26) considered that information, and thus behavioral response, may only be available locally rather than globally and Weitz et al. (20) considered that behavioral response may react to the incidence of mortality rather than infection. The general extension of the standard modeling framework for infectious diseases that we have proposed offers a pathway to guide more specific mechanistic investigations.

The description of these new equilibria represents a novel advance for infectious disease and vaccination policy development. A stable equilibrium provides a policy target where the system is self-managing and self-maintaining. The existence of such a stable target allows optimal policy strategies to be formulated to reach that point. Policy formulation without such an explicit goal requires iterative trial-and-error which may incur economic or societal costs that can undermine support for the process. Such adaptive control strategies built upon iterative learning have a long history (36–38) and are useful tools complemented by our results showing that there are multiple advantageous stable equilibria (disease-free or endemic) allowing a return to normal behavior. In the face of uncertainty about the feasibility of elimination, the endemic state with return to normal behavior provides a valuable new policy target to motivate action and guide policy development.

## Materials and Methods

We will demonstrate the power of of our approach on the most basic infectious disease model. Thus, we start with the Susceptible, *S*, Infected, *I*, Recovered, *R*, (SIR) model for a well-mixed population given by,

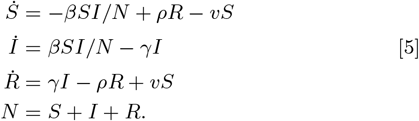

with transmission rate *β*, average duration of illness 1*/γ*, and a conserved population *N*. The parameter, *v*, represents the vaccination rate, which moves population from susceptible, *S*, to recovered, *R*. Conversely, the parameter, *ρ*, represents loss of immunity, which moves population from recovered, *R*, back to susceptible, *S*. Note that this model for vaccination implicitly includes booster immunizations, since loss of immunity will eventually move the previously vaccinated population back into the susceptible class and the model assumes that they may eventually be re-vaccinated or “boosted”. The specific interpretations of the terms and parameters in Eq. (5) is provided only for aiding in intuition. For example, instead of reinfection the source of new susceptible population may be births (on a longer time scale), or there may be other methods of removing people from the susceptible population besides vaccination. The model and analysis presented here may be adaptable to such interpretations since our focus will be on the interaction of behavior and transmission.

The key to a frequency-based transmission model such as Eq. (5) is the nonlinear (*S* multiplies *I*) term for case incidence,

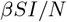

which quantifies the incidence rate of the disease. Following (39), we can break down the transmission rate, *β*, into a product of the rate of effective contact, *a*, and the probability of transmission given an effective contact, *B*, so that,

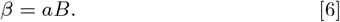

The *activity* rate, *a*, represents an *effective* rate that can include changes in behavior such as distancing or masking. To see this, it is worthwhile to note the formal descriptions of *a* and *B* from (39): *a* represents the rate of contacts that are of an appropriate type for transmission to be possible if one of the hosts is infectious, and *B* represents the probability that contact between an infectious and a susceptible host does in fact lead to transmission. Using these definitions, this framework allows for activity change that both reduces the rate of all contacts (e.g. distancing) or the rate of effective contacts (e.g. masking).

Substituting Eq. (6) into the formula for case incidence, *C*, we define,

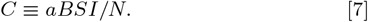

In this product *S* is the susceptible population who are having effective interactions with people at rate *a*. The probability of each interaction happening with an infected person is *I/N*, and *B* is the conditional probability that such an interaction with an infected person gives rise to infection. By separating *β* into its component factors, we see that it is much more reasonable to assume that *B* is constant (or at least that it changes on a longer time scale), whereas a behavioral response could be quite rapid and makes it likely that the rate of effective contact, *a*, could change on fast time scales. Notice that when *a* = 1 we have *β* = *B* so we refer to *a* = 1 as the *baseline* level of activity.

We are now ready to quantify the various assumptions (A1-A3) that we will consider for the behavioral dynamics. First, *Reactivity* (Assumption A1) says that the rate of change of the activity parameter is a continuous function, *F*, that only depends on the current activity, *a*, and case incidence rate, *c*, which is the rate of new infections, *c*≡*C/N* (here *C* is raw incidence and *c* is the incidence as a percentage of the total population). In other words, reactivity allows any behavioral dynamics of the form,

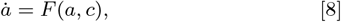

and we call *F* the *reactivity function*. The fact that there cannot be a ‘negative’ infection incidence implies that *a*≥ 0. When *a* = 0 the rate of change of activity cannot be negative. Thus, in addition to the form Eq. (8), reactivity also includes the assumption that *F* (0, *c*) ≥0.

We can now quantify *Resilience* (Assumption A2), which states that when incidence is zero activity will increase. Here we come to one of the significant advantages of not specifying a model for activity. Recall that the baseline activity level is defined to be the level of activity that would be reached if the disease were removed and a long time were allowed for the activity to stabilize. Since the reactivity function, *F*, is not specified, we can always choose units for *a* such that the baseline activity level is *a* = 1 by incorporating the change of units into the definition of the reactivity function.

When *a* = 1, transmissibility during contacts reflects the baseline contagiousness of the disease. All we are assuming here is that there is *some* baseline value for activity, and then choosing units which re-scale that value to one. Thus, without loss of generality, resilience can be quantified as,

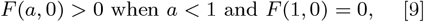

which implies that when activity is below baseline activity (*a* < 1) and incidence is zero (*c* = 0) activity will increase 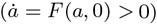. We also need to assume that *F* (1, 0) = 0 to insure that baseline activity is stationary when there is no incidence. The condition Eq. (9) is all that is required when we are also assuming boundedness (A3), but for technical reasons when the behavior is not bounded we will also assume *F* (*a*, 0) < 0 when *a* > 1. Note that we have assumed we are working in units where *a* = 1 corresponds to baseline activity, so *a* < 1 means any level of activity that is below baseline and *a* > 1 means activity is above baseline. Moreover, *F* (*a*, 0) > 0 means that, when there is no incidence, the rate of change of activity is positive, so activity is increasing, and this captures the assumption of resilience. Resilience also includes the assumption that baseline activity (*a* = 1) is stationary when there is zero incidence (*c* = 0); this assumption is captured by the equation *F* (1, 0) = 0 in Eq. (9).

Lastly, *Boundedness* (Assumption A3) says that the baseline activity level (averaged over the whole population) is the highest level possible, meaning that *a*≤1. This means that when *a* = 1 we must have

(A3 : Boundedness)

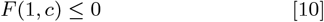

otherwise the activity would increase beyond the boundedness limit of *a* = 1.

Thus, we consider the following infection model that incorporates vaccination, loss of immunity, and arbitrary behavioral dynamics,

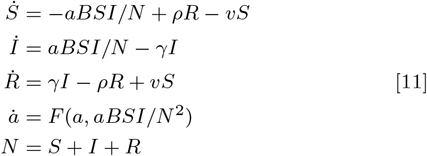

In order to remove the algebraic equation *N* = *S* + *I* + *R* we rewrite the model in terms of population fractions. Setting *s* = *S/N, i* = *I/N*, and *r* = *R/N* we have *s* + *i* + *r* = 1 and 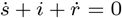. Moreover, we can remove the equation for the recovered population fraction, *r*, by setting *r* = 1−*s*−*i* in the remaining equations. Thus, the following equations govern the fractions of the population,

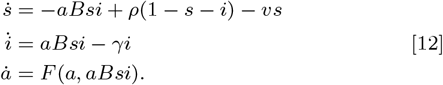

In these units, the basic reproduction number is *R*_0_ ≡*B/γ*, which defines the expected number of secondary infections due to the initial infection in a completely naive population.

## Data Availability

All data produced in the present work are contained in the manuscript or available online at https://github.com/CSSEGISandData/COVID-19

## ACKNOWLEDGMENTS

Supported by US NIH Director’s Transformative Award 1R01AI145057.

## S. Supporting Information: Stabilizing the return to normal behavior in an epidemic

### S.1. Theorems and Proofs

We first show that the disease dynamics are contained in the region shown in Fig. 2 and can never leave. Under the assumptions A1-A3, there is a region, Ω, (Fig. 2), such that if the system starts within Ω, then the system will remain inside Ω at all future times (Lemma 1 below). This property ensures that no populations become negative and that the activity variable *a* stays within its prescribed range 0≤*a*≤1. Thus, all long-term behavior will be determined by dynamical attractors which could be equilibria, cycles, or even chaos (40), which must lie entirely within Ω.

#### Lemma 1.

*Under the dynamics of* Eq. (12), *the set*

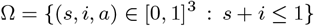

*is invariant when F is resilient* Eq. (9) *and bounded* Eq. (10).

*Proof*. Note that when *a* = 0 we have *F* (0, 0) ≥0 by Eq. (9) (resilience), and when *a* = 1 we have *F* (1, *Bsi*) ≤0 by Eq. (10) (boundedness). So the *a* component is always pointing into Ω and we need only consider the (*s, i*) variables. When 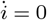 we have *i* = 0 and when 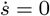 we have 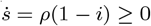, so along each of these boundaries the vector field is pointing into the set Ω. Finally, we check the boundary *s* + *i* = 1, where 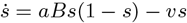 and 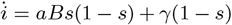 so that 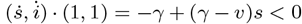 meaning that the vector field is always pointing into the set (since (1,1) is the outward pointing normal vector to the boundary *s*+*i* = 1 and the inner product of the vector field with this outward pointing normal is negative). □

The lemma shows that for any reactive behavioral response function, *F*, which is resilient and bounded, the dynamics of the disease will always preserve some natural constraints that we expect. For example, the variables *s, i*, and *r* represent fractions of the population and so they should always be between zero and one and they should always sum to one. If we view the variables *s* and *i* as lying in a plane, these constraints imply that they must always lie in a triangle as shown in Fig. S.1. If we now add the activity variable in the vertical dimension (coming out of the plane of the paper in Fig. S.1), we see that we have a solid shape with horizontal triangular cross sections as shown in Fig. 2.

Another natural limitation is that the activity variable, *a*, cannot be negative, since that would imply that members of the infected population are moving directly back into the susceptible population. The model does allow loss of immunity and re-infection through the *ρ* parameter, but as an axiom we do not permit ‘negative’ infections. It would also be possible to allow immediate re-infection by using a term proportional to the recovered and infected populations, however, for simplicity we assume here that recovery imputes at least a temporary immunity, and the length of time of this immunity is controlled by *ρ*. Altogether, taking the solid shape with triangular horizontal cross-sections and restricting the height to be between zero and one we have the gray solid shape shown in Fig. 2 which we call the *domain*, Ω, of the dynamics.

The way we constructed the domain in Fig. 2, the dynamics of the disease *should* be constrained to that region for all time, however, a poorly specified dynamical model could allow the dynamics to ‘escape’ the domain, violating our axioms. So our first result is Lemma 1, which shows that for any resilient and bounded activity function, the state cannot escape and is confined to the domain in Fig. 2 forever. This result, although a bit cumbersome to check, simply requires showing that along each surface of the solid shape all the arrows of the dynamics are always pointing inwards. Notice that our key assumptions of resilience and boundedness concern the boundaries of the domain. For example, resilience says that *F* (*a*, 0) > 0, meaning that it only constrains what happens when the rate of incidence, *c* = *aBsi*, is zero, which is only true when either *i* = 0 or *s* = 0 or *a* = 0, and these are the front square surface, left square surface, and bottom triangular surface of the domain respectively. Resilience says that along each of these three surfaces, the vertical component of the arrows that define the dynamics are pointing upwards, towards increasing activity. Boundedness says that *F* (1, *c*) ≤ 0, and activity is only equal to along the top triangular surface of the domain. So boundedness implies that along this top surface the vertical component of the arrows that define the dynamics must not point up (they can point down or horizontally or be zero).

Our main result on equilibria is stated next, and will be derived in stages.

**Fig. S.1.**
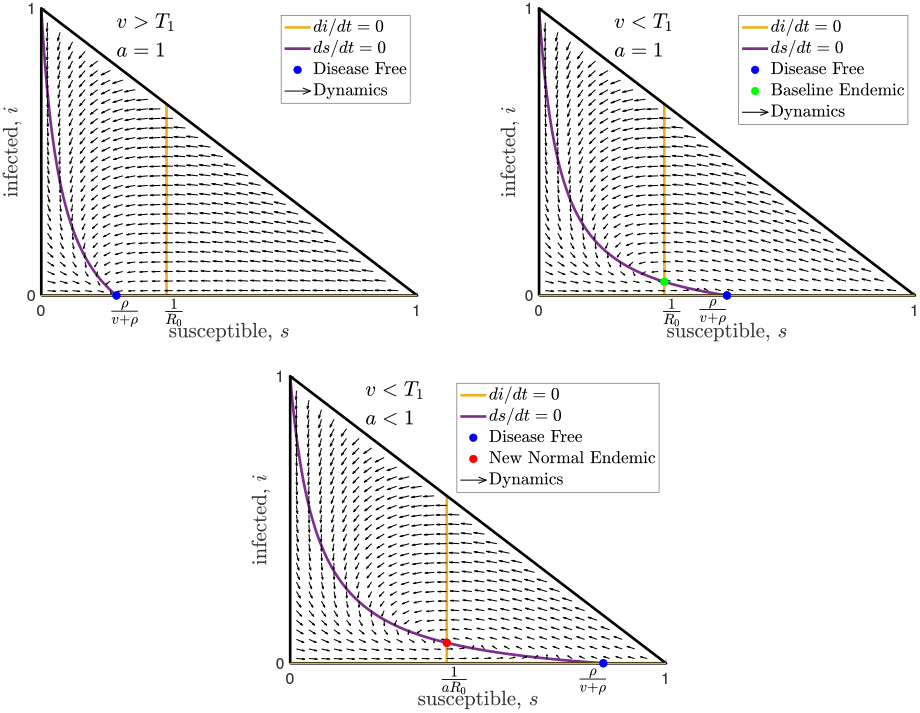
Equilibria and dynamics projected on the susceptible/infected plane. Here we show three horizontal slices (fixed activity slices) from the domain, Ω, shown in Fig. 2. The top left and top right slices show the top of the domain (*a* = 1) with high and moderate vaccination rates respectively. The bottom slice is from the middle of the domain when the vaccination rate is very low and only the new normal endemic is stable. Note that the flow arrows, curves, and intersections shown in these cross sections depend on the disease parameters but are independent of the behavioral dynamics (except for the activity level, *a*, of the new normal endemic shown in the bottom panel).

#### Theorem 2.

*Consider the reactive dynamics* Eq. (12) *for any twice differentiable F* : [0, 1]^2^ →ℝ *that is both resilient and bounded. Then*,

- *There is only one disease-free equilibrium, and it has baseline activity (a* = 1).
- *When the vaccination rate is high enough, namely*,

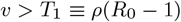

*the disease-free equilibrium is the only equilibrium in* Ω *and it is locally asymptotically stable*.
- *When v* < *T*_1_ *the disease-free equilibrium is unstable and there exists a unique baseline endemic equilibrium (a* = 1*) and at least one new normal endemic equilibrium* (*a* < 1).
- *The baseline endemic equilibrium is stable when*,

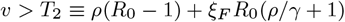
- *where ξ*_*F*_ *is a constant that depends on F*.
- *If the vaccination rate is below T*_2_ *then the only stable equilibria are new normal endemic equilibria (a* < 1*) and at least one new normal endemic equilibrium must exist*.

Theorem 2 follows immediately from Theorems 3-6 below. For the *T*_2_ threshold, we do not have an explicit formula for the constant *ξ*_*F*_, however we will show that it is given by 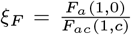 for some *c* ∈(0, *γi*), and we will show that *F*_*a*_(1, 0) < 0 so when *F*_*ac*_(1, *c*) > 0 we have *T*_2_ < *T*_1_. It is also possible to have *T*_2_≥*T*_1_ and in these cases the baseline endemic is never stable. In practice, it is straightforward to find the second vaccination threshold by solving the equation 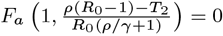 for *T*_2_. For examples that show how to find *T*_2_ see Section S.2.

To motivate the basic dichotomy for equilibria in terms of infection rate (disease-free vs. endemic), note that setting *di/dt* = 0 immediately implies that either *i* = 0 or *aBs* − *γ* = 0. The former case is the disease-free equilibrium, and, in the latter case, one can show that the fraction of the infected population will be,

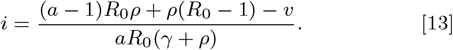

This cannot be negative, and since *a*≤1 the first term in the numerator is negative or zero, so we immediately see that when the vaccination rate is greater than *ρ*(*R*_0_−1) there cannot be any equilibria of the form Eq. (13). At this point we have only made Assumption A1, and we already have a universal vaccination threshold which we call,

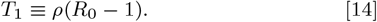

When the vaccination rate is above *T*_1_, the only possible equilibrium is the disease-free equilibrium, and this holds for any reactivity function *F*.

In order to analyze the stability of equilibria, we will frequently make use of the Jacobian matrix of partial derivatives of the right hand side of Eq. (12) which is,

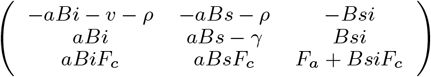

where *F*_*a*_ and *F*_*c*_ are shorthand for partial derivatives and are evaluated at (*a, aBsi*) in the Jacobian.

#### Theorem 3

(Disease-Free Equilibrium). *For any reactive dynamics of the form* Eq. (12), *every disease-free equilibrium has* 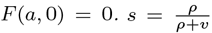 *and the equilibrium activity level solves F* (*a*, 0) = 0. *When the vaccination rate, v, is below the universal threshold, T*_1_≡*ρ*(*R*_0_ −1) *all disease-free equilibria are unstable*.

*If we also assume that the behavior is resilient, there is only one disease-free equilibrium. It has baseline activity (a* = 1*), and is stable when the vaccination rate is greater than T*_1_.

*Proof*. Setting the equations in Eq. (12) equal to zero and substituting *i* = 0 for a disease-free equilibrium we immediately find that *B* = *b* = 0 and 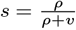 and *F* (*a*, 0) = 0. The Jacobian of the right hand side of Eq. (12) at this equilibrium is,

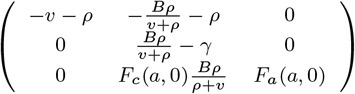

and the eigenvalues are *λ*_1_ = −*v* − *ρ*, 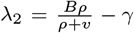, and *λ*_3_ = *F*_*a*_(*a*, 0). When *v* < *T*_1_ we have *λ*_2_ > 0 which implies that the equilibrium is unstable. Since this Jacobian applies to any disease-free equilibrium, this means that a vaccination rate below *T*_1_ implies that any disease-free equilibrium will be unstable.

If we assume the behavioral dynamics are resilient, we have immediately that *F* (1, 0) = 0 so that there is a disease-free equilibrium with baseline activity, *a* = 1. Moreover, resilience says that *F* (*a*, 0) > 0 whenever *a* < 1, so there is only one disease-free equilibrium, and it has baseline activity. To analyze the stability of this disease-free equilibrium, note that *F* (1, 0) = 0 implies

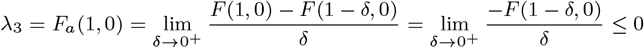

where the final inequality follows from the fact that *F* (1−*δ*, 0) > 0 by resilience. So when *F*_*a*_(1, 0) < 0 and *v* > *T*_1_, all the eigenvalues of the Jacobian are negative and the disease-free equilibrium is asymptotically stable. The cases of *v* = *T*_1_ and *F*_*a*_(1, 0) = 0 are special cases known as non-hyperbolic equilibria, and stability in these cases have to be determined separately from the general stability analysis. Since it is unlikely that the vaccination rate would be exactly equal to *T*_1_, we will leave that case aside (which is why we assume *v* is strictly less than *T*_1_ in the statement of the result). However, to avoid additional assumptions on *F*, we must also consider the non-hyperbolic case when *F*_*a*_(1, 0) = 0.

While the stability of non-hyperbolic equilibria is typically difficult to analyze, in the case of a bounded behavioral model we can prove that the disease-free equilibrium is still stable even when *F*_*a*_(1, 0) = 0 (as long as *v* > *T*_1_). To show this, we will use a technical notion of stability known as Lyapunov stability, which says that for any small region around the equilibrium, we can find an even smaller region such that if you start in the smaller region you will never leave the first region. We will prove this by finding ϵ_0_ > 0 such that for any ϵ ∈ (0, ϵ_0_) we can find an invariant open set inside the ϵ-ball that contains the equilibrium. The basic intuition is that because the system has two negative eigenvalues, when we get close enough to the equilibrium, the first two components of the vector field will be pointing into a cylinder surrounding the equilibrium. Moreover, because of resilience, the vector field must be pointing upwards on the bottom of the cylinder (as long as the cylinder is taken sufficiently small), and assuming boundedness the dynamics cannot leave the top of cylinder. Thus, the cylinder will be invariant under the dynamics.

Fix ϵ > 0, then for each *a* ∈ (1−ϵ, 1) we have *F* (*a*, 0) > 0 by resilience, and since *F* is continuous there must exist some *δ*_*a*_ such that *F* (*a, c*) > 0 for all *c* < *δ*_*a*_ and we can choose *δ*_*a*_ to be a continuous function of *a*. Let *δ* = max_*a*∈ [1−ϵ,ϵ]_ {min {ϵ− (1−*a*), *δ*_*a*_}}, which exists since it is a continuous function on a compact set, and let *ā* be the largest value of *a* with *δ*_*a*_ = *δ*. We can now define a cylindrical region 𝒪 = {(*s, i, a*) : ||(*s, i*) − (*ρ/*(*ρ* + *v*), 0) ||< *δ/* max {1, *B*}, |*a*−1| < 1−*ā*}. To see that 𝒪 is invariant, note that the bottom of the cylinder is the set with *a* = *ā* and *δ*_*ā*_ = *δ*, so *F* (*ā, c*) > 0 for all *c* < *δ* and || (*sρ/*(*ρ* + *v*), *i*) || < *δ/B* so we have *c* = *aBsi*≤*B*||(*s, i*) − (, 0)| < *δ* (since *a, s, ρ/*(*ρ* + *v*) ≤1 and *i* < *δ/B*) so each point on the bottom of the cylinder is within the radius where 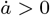, so the vector field is pointing up into the cylinder. Note that a similar argument can be made for the top of the cylinder using the fact that resilience includes *F* (*a*, 0) < 0 when *a* > 1, or simply using boundedness in which case the top of the cylinder is just *a* = 1.

Next we need to show that the vector field along the walls of the cylinder is pointing into the cylinder, and this will require taking ϵ > 0 sufficiently small. We first use Taylor’s theorem to argue that sufficiently close the equilibrium the vector field looks close to its linearization namely, writing Eq. (12) as 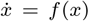 where *x* = (*s, i, a*)^T^ and calling the equilibrium *x*_0_ = (*s, i, a*)^T^ = (*ρ/*(*ρ* + *v*), 0, 1)^T^ we have, *f* (*x*_0_) = 0 so Taylor’s theorem says,

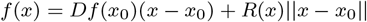

where *R*(*x*) →0 as *x*→*x*_0_. Now take the vector *x*−*x*_0_ which points away from the equilibrium and orthogonally decompose it as *x*−*x*_0_ = *v* + *v*^*a*^, where *v*^*a*^ is the component in the *a*-direction and *v* is the projection of *x*−*x*_0_ into the (*s, i*)-plane. Taking the inner product *v* we have

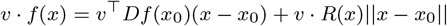

and in the non-hyperbolic case

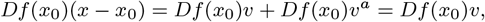

so writing 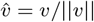 we have,

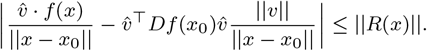

Now note that along the walls of the cylinder O we have 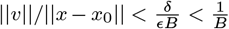(since *δ* < ϵ), so choose ϵ_0_ > 0 sufficiently small so that for all ||*x*−*x*_0_|| < ϵ we have ||*R*(*x*) || < max {*λ*_1_, *λ*_2_}. Then 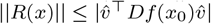 (since the latter is the Raleigh quotient for *Df* (*x*_0_) orthogonal to (0, 0, 1)^T^ and *λ*_1_, *λ*_2_ < 0 are its eigenvalues in that subspace) and it follows that 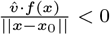 (since *R*(*x*) is sufficiently small that it must have the same sign as 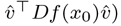. Notice that 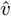 is the orthogonal projection into the (*s, i*)-plane of the vector pointing away from the equilibrium (and is thus normal to the cylinder wall pointing outwards). Thus the vector field *f* (*x*) must be pointing into the cylinder𝒪, since its inner product with 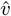 is negative.

So we conclude that for resilient behavior with *F*_*a*_(1, 0) < 0 the disease-free equilibrium is asymptotically stable, and even when *F*_*a*_(1, 0) = 0 the disease-free equilibrium is Lyapunov stable. □

Thus we have seen that the disease-free equilibrium requires stabilizing baseline activity (*a* = 1), and for resilient behavioral dynamics a sufficient condition is a high enough vaccination rate, *v* > *T*_1_.

Next we turn to equilibria that have nonzero disease levels. Assume there is an equilibrium (*s, i, a*) of Eq. (12) with *i* > 0. Solving for the equilibrium yields

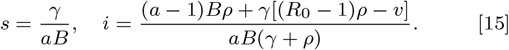

Notice that since *a*≤1, having a vaccination rate *v* > *ρ*(*R*_0_−1) implies that *i* < 0, so no endemic equilibrium exists. Recall that this parameter range is exactly where the disease-free equilibrium is stable (Theorem 2).

On the other hand, if *v* < *ρ*(*R*_0_−1), then for every value of *a* satisfying *F* (*a, c*) = 0 an endemic equilibrium (*s, i, a*) exists. In other words, these endemic equilibria are created as the vaccination rate *v* drops through the stability threshold *T*_1_ = *ρ*(*R*_0_ − 1).

#### Theorem 4

(Baseline Endemic Equilibrium). *For any F the model* Eq. (12) *has at most two equilibria with baseline activity (a* = 1*) in* Ω, *namely, the disease-free equilibrium from Theorem 3, and an endemic equilibrium with* 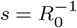 *and* 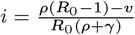*This second equilibrium is called the baseline endemic equilibrium, it exists when F* (1, *γi*) = 0, *and it is only in* Ω *when v* < *ρ*(*R*_0_−1) *(meaning the disease-free equilibrium is unstable). When F is Bounded (satisfies* Eq. (10)*), this endemic equilibrium is stable if and only if F*_*a*_(1, *γi*) ≤ 0. *Moreover, there exists c*∈(0, *γi*) *such that the stability condition is*,

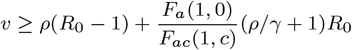

*and the equilibrium is locally asymptotically stable if the inequality is strict*.

*Proof*. Setting *a* = 1 the equation 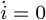 immediately reveals that either *i* = 0 (the disease-free equilibrium) or 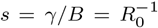. Setting 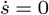 we find 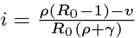 and the instantaneous case rate is *C/N* = *Bsi* = *γi*. Now, by Eq. (10), for any *δ* > 0 we have, *F* (1, *γi* + *δ*) ≤0 and *F* (1, *γi* −*δ*) ≤ 0, and since we are at equilibrium we have *F* (1, *γi*) = 0 which implies

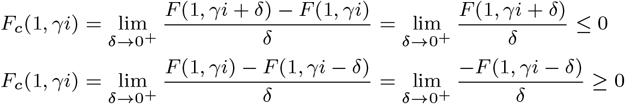

so we have *F*_*c*_(1, *γi*) = 0. This fact simplifies the Jacobian at the equilibrium which becomes

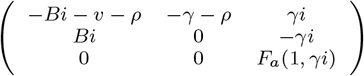

with characteristic equation

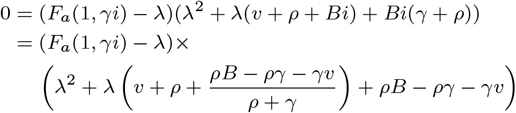

Thus, *λ* = *F*_*a*_(1, *γi*) is an eigenvalue and the remaining two eigenvalues are

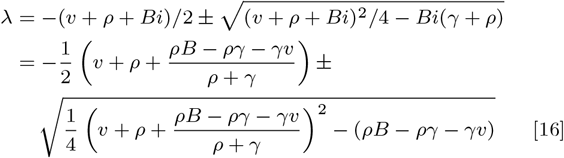

since all these variables and parameters are positive, from the first expression for *λ* we have

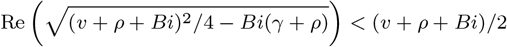

so that the real part of both of these eigenvalues are negative. Thus when *F*_*a*_(1, *γi*) > 0 this equilibrium is unstable and when *F*_*a*_(1, *γi*) < 0 it is stable. Now by the mean value theorem, there exists *c* ∈ (0, *γi*) such that,

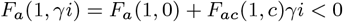

and plugging in for *i* and solving for the vaccination rate we find 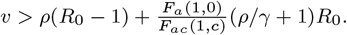

Recall that *F*_*a*_(1, 0) ≤0, so assuming *F*_*ac*_(1, *c*) > 0, Theorem 4 establishes the existence of a lower vaccination threshold for stabilizing the baseline endemic equilibrium. Now we turn to the existence of a new normal (*a* ≠1) endemic (*i* ≠ 0) equilibrium.

Notice that in the proof of Theorem 4 there is the possibility of complex eigenvalues meaning that the dynamics near the baseline endemic equilibrium would behave like a damped harmonic oscillator. However, this oscillatory behavior is entirely due to the SIR dynamics, and does not arise from the behavioral dynamics. Behavioral oscillations are also possible, but will depend on the specific form of the *F* function that defines the behavioral dynamics. Moreover, behavioral oscillations will only arise from the new normal endemic equilibria addressed in Theorem 6 below. Before considering new normal endemic equilibria, we first characterize the possible oscillations of the baseline endemic equilibrium.

#### Corollary 5.

*The dynamics near the baseline endemic equilibrium from Theorem 4 will be oscillatory when B is in the range*

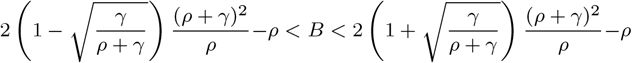

*and the vaccination rate is sufficiently low, namely*

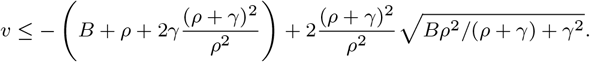

*The damping rate and frequency of the oscillation near the equilibrium are given by*,

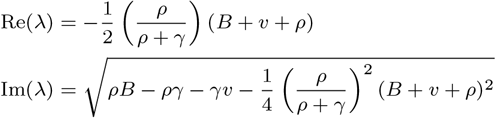

*(where λ is the complex eigenvalue from the proof of Theorem 4) and the damping ratio of the oscillation near the equilibrium is*,

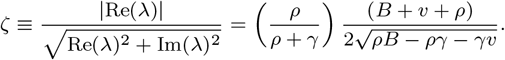

The proof of Corollary 5 follows directly from the formulae for the eigenvalues in Theorem 4 by elementary (albeit cumbersome) algebra and is omitted.

#### Theorem 6

(“New Normal” Endemic Equilibria). *Assume that v* < *ρ*(*R*_0_−1) *(which implies that the disease-free equilibrium is unstable) and that the baseline endemic equilibrium is unstable. For any F that is Resilient and Bounded (meaning F satisfies* Eq. (9) *and* Eq. (10)*) the dynamics of* Eq. (12) *have at least one new normal endemic equilibrium in* Ω *with a* ≠ 1 *and i* ≠ 0. *Conversely, when v*≥*ρ*(*R*_0_−1) *there are no new normal endemic equilibria in* Ω.

*Proof*. Since we are assuming *i* ≠0 and *a* ≠1, setting 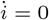 we find 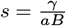 and plugging this into 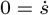 we find 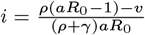. Thus, at any equilibrium we can write

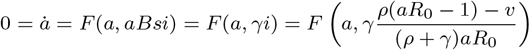

so we efine a curve Γ : [0, 1]→ ℝ^2^ given by 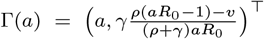. The endpoints of the curve are 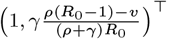 and (0, −∞)^T^, so as *a* → 0 the curve leaves [0, 1]^2^ and hits the boundary when 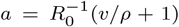. Thus, the interval of *a*-values for which the curve is in Ω is 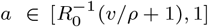. Note that if *ρ*(*R*_0_−1) < *v* then the left hand endpoint is greater than 1, so the curve never enters [0, 1]^2^ and there are no new normal equilibria. Otherwise, at the left endpoint, we have 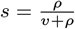 and *i* = 0, so we have 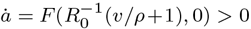 by Resilience. At the other endpoint, we have the baseline endemic equilibrium with *a* = 1 and 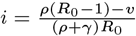 and 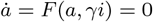.

Moreover,

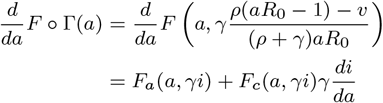

and, since *F* is bounded, at the baseline equilibrium we have,

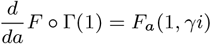

since *F*_*c*_(1, *γi*) = 0 as shown in Theorem 4. Recall from Theorem 4 that the baseline equilibrium is stable when *F*_*a*_(1, *γi*) < 0 and unstable when *F*_*a*_(1, *γi*) > 0. Thus, when the baseline equilibrium is unstable, we have *F*_*a*_(1, *γi*) > 0, so 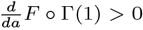, so for all *â* < 1 sufficiently close to 1 we will have *F* ° Γ(*â*) < 0. Now by the intermediate value theorem, there must be an *a*^*^ between 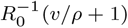 and *â* such that *F* ° Γ(*a*^*^) = 0 and this is a new normal equilibrium. □

### S.2. Analysis of Examples

In this section we show how to apply the Theorems above to analyze reactivity functions using the examples from Section 2.

#### Example 1: Linear Response

The first reactivity function we consider is the linear model Eq. (2) given by

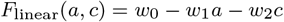

note that 0 = *F* (1, 0) = *w*_0_−*w*_1_, so *w*_0_ = *w*_1_ and we can rewrite this model as,

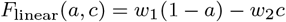

where *w*_1_, *w*_2_ > 0 so that when *a* < 1 we have *F*_linear_(*a*, 0) = *w*_1_(1−*a*) > 0 so *F*_linear_ satisfies resilience, and *F*_linear_(1, *c*) =−*w*_2_*c*≤ 0 so *F*_linear_ satisfies boundedness. At any endemic equilibrium we have *aBs* = *γ* so the incidence is give by

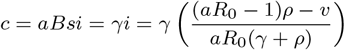

where we used an alternate form of Eq. (13) for *i* at equilibrium. Substituting this into *F*_linear_ and setting equal to zero to find the equilibrium we have,

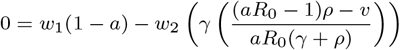

so

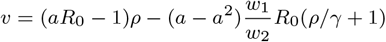

Notice that for *F*_linear_ there is a quadratic relationship between vaccination and activity at equilibrium, as illustrated in Fig. 3a. However, *F*_linear_ does not necessarily have a baseline endemic equilibrium since setting *F*_linear_(1, *c*) =−*w*_2_*c* = 0 implies the only solution is *c* = 0, which is the disease-free equilibrium. Note that if we set *w*_2_ = 0 then every *c* solves *F*_linear_(1, *c*) = 0 so there are many baseline endemic equilibria, and the stability condition is *F*_*a*_(1, *γi*) = −*w*_1_

#### Example 2: Quadratic Response

The second reactivity function we consider is the quadratic model Eq. (3), given by

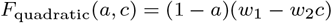

where *w*_1_, *w*_2_ > 0. If we substitute *c* = *aBsi* we see that this model is quadratic in activity, *a*. Notice that when *a* < 1 we have *F*_quadratic_(*a*, 0) = *w*_1_(1 − *a*) > 0 so *F*_quadratic_ satisfies resilience, and *F*_quadratic_(1, *c*) = 0≤0 so *F*_quadratic_ satisfies boundedness. At any endemic equilibrium we have *aBs* = *γ* so the incidence is give by

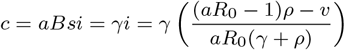

where we used an alternate form of Eq. (13) for *i* at equilibrium. Substituting this into *F*_quadratic_ and setting equal to zero to find the equilibrium we have,

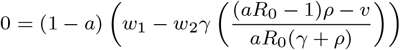

so either *a* = 1 (the baseline activity) or we have a new normal endemic equilibrium at

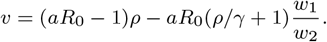

Notice that for *F*_quadratic_ there is a linear relationship between vaccination and activity as illustrated in Fig. 3a. To solve for the second vaccination threshold, *T*_2_, we set, *F*_*a*_(1, *γi*) = 0, and solve for the vaccination rate, *v*. For *F*_quadratic_ we have,

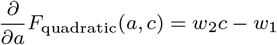

so that 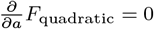 implies

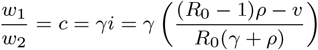

and solving for *v* gives the second vaccination threshold,

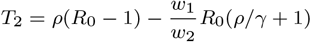

#### Example 3: Bilinear Response

The third reactivity function we consider is the bilinear function Eq. (4), given by

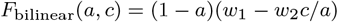

where *w*_1_ > 0 and *w*_2_ > 0. It is easy to see that *F*_bilinear_ satisfies resilience and boundedness. If we rewrite this model in terms of activity and infection rate we have,

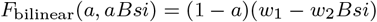

so that this model is bilinear in activity and infection rate (rather than incidence). To solve for the equilibrium, we compute,

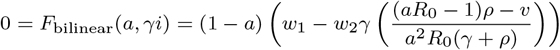

so we have an baseline solution, *a* = 1, and a new normal solution,

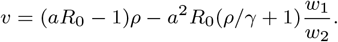

To find the second vaccination threshold we set

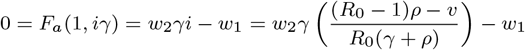

and solve for *v* to find,

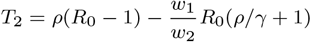

The connection of 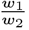 with the threshold *T*_2_ suggests that this ratio (and its reciprocal 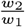) determine the strength of the response. In Fig. S.2 we show that when 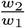 is small (top row, yellow curves) the behavioral response is weak and dynamics approaches that of a classical SIRS model. Moreover, when 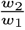 is large, the behavioral response is more robust and can even lead to an increasing series of waves. We should note while the bilinear response can create an arbitrarily long series waves with almost equal peaks (as shown in the next section) this particular response function requires *v* > 0 to obtain increasing waves. It may be worth considering that a small *v* > 0 could be used to model a subset of the population that become infected and recover without ever becoming *infectious*, and thus never entering the *i* class.

Such a small percentage of cases could potentially arise from very mild infections or through extremely rigid isolation that removes the possibility of infecting others entirely. Of course, if one is interested in capturing the increasing waves observed in Fig. 1, one could also consider other response functions and we have empirically observed increasing waves without vaccination using more complicated response functions.

**Fig. S.2.**
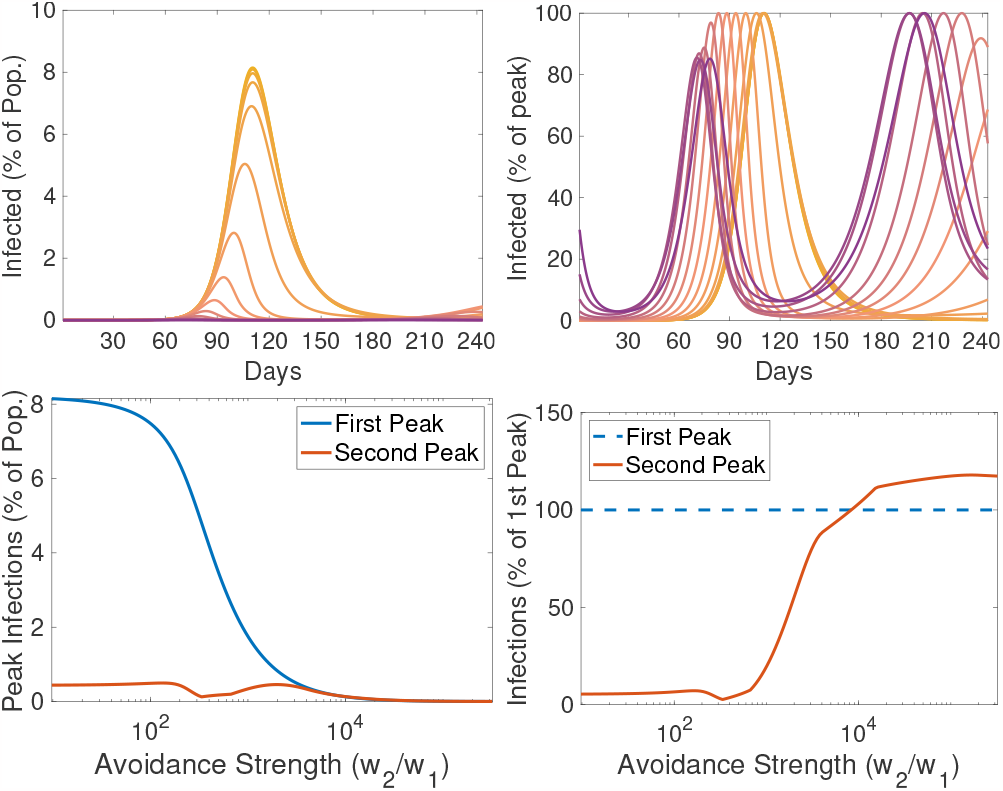
Top: Time series of infectious population size with various levels of avoidance strength (*w*_2_*/w*_1_) colored in a gradient from weak (yellow) to strong (purple). Infectious population size is shown in terms of percentage of population (left) and in terms of percentage of the peak infections over the time window (right), mirroring Fig. 1 from the manuscript. Bottom: In the bilinear response, peak infections decay quickly with increasing avoidance strength (left), while the height of the second increases as a percentage of the first peak height until it eventually exceeds the first peak (right) which is not observed in the classical SIRS model.

### S.3. Connection to Predator-Prey Models

If we combine the reactivity function *F*_bilinear_ from Eq. (4) with the framework Eq. (12) we have the example model,

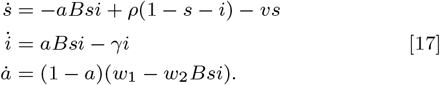

In order to understand the early epidemic dynamics of this model, consider *s*≈1 as approximately constant and set *d* = 1−*a* so that Eq. (17) can be approximately reduced to,

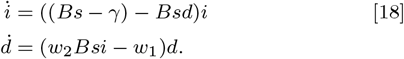

which is exactly the Lotka-Volterra predator-prey model. Perhaps counter-intuitively, in this analogy the infections play the role of prey and ‘distancing’, *d*, plays the role of the predator.

Regardless of the analogy, the interesting feature of this model is that it produces oscillations with frequency 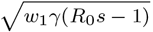. This approximation is valid when these activity driven oscillations are fast enough that *s* is approximately constant over the course of an oscillation. Each oscillation reduces *s* slightly, and over time the frequency decreases. Eventually, when 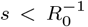, we have *R*_0_*s*−1 < 0 which changes the stability of the equilibrium inside the periodic orbit of Eq. (18) from a center to a source. Thus, 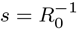 represents a phase transition threshold for this model.

In Fig. 4 we illustrate the range of dynamics that this simple reactivity function can exhibit. In this example the infectiousness period is 6 days, loss of immunity is 300 days, the infectiousness parameter is *B* = 0.5, the reactivity function parameters are *w*_1_ = 0.01 and *w*_2_ = 100. In the high vaccination case (Fig. 4a), the system passes through the phase transition quickly and the dynamics resemble a classical epidemic. Similarly, when the vaccination rate is moderate (Fig. 4b) the system quickly relaxes to the baseline endemic equilibrium. In the low vaccination case (Fig. 4c) the oscillations continue for an extremely long time (in fact they are very slowly decreasing in amplitude but would still be visible after 100 years). In Fig. S.3 we show that by increasing the reactivity to *w*_1_ = 0.035, the high and moderate vaccination dynamics can initially exhibit predator-prey type oscillations until the susceptible population is reduced to the phase transition level, 1*/R*_0_, at which point the oscillations become classically damped.

**Fig. S.3.**
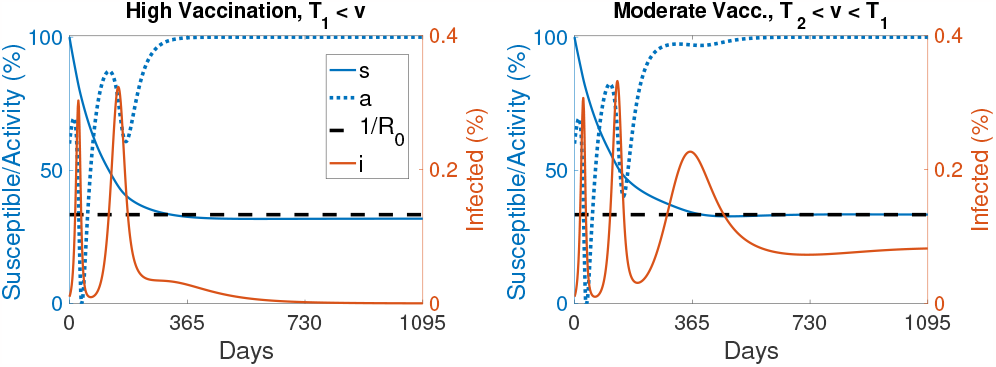
Increased reactivity leads to oscillations even with high and moderate vaccination. Repeating the simulations from the top row of Fig. 4 but increasing the value of the *w*_1_ parameter to *w*_1_ = 0.035.

